# A Psychometric Examination of the Dimensional Obsessive Compulsive Scale in a Treatment-Seeking Youth Sample

**DOI:** 10.1101/2024.09.06.24313128

**Authors:** Nicholas R. Farrell, Catherine W. MacDonald, Mia Nuñez, Andreas Rhode, Nicholas Lume, Patrick B. McGrath, Marina Baskova, Eli Wilson, Jonathan S. Abramowitz, Jamie D. Feusner

## Abstract

The Dimensional Obsessive Compulsive Scale (DOCS) was developed to address several limitations of existing self-report measures of obsessive compulsive disorder (OCD) symptoms, and has been found to be a psychometrically sound method of assessing OCD symptoms in adults. However, to date, the psychometric performance of the DOCS has not been studied in a youth sample. The present study addressed this gap in the literature by examining the psychometric properties of the DOCS in a large sample (n=182) of treatment-seeking youth diagnosed with OCD. Results indicated that the DOCS showed good convergent validity with a youth OCD assessment scale, as well as similar sensitivity to the effects of treatment-related change in symptom severity. The DOCS also maintained its original four-factor structure in the youth sample, similar to findings in adults, supporting the consistency of the four subscales included. Overall, the DOCS appears to represent a promising method for assessing OCD symptom severity and response to treatment of OCD in youth.

## Introduction

Obsessive-compulsive disorder (OCD) is a chronic and highly debilitating mental health condition that is responsible for significant disability in affected individuals (Mancebo et al., 2008) as well as considerable economic burden on society (Kochar et al., 2023). Although OCD can occur at any age across the lifespan, it appears to be particularly prevalent in youth. Indeed, research has consistently shown that one of the peak times of OCD onset is around age 10 (Geller et al., 2006). OCD is responsible for a myriad of unique problems in youth, such as school refusal, academic deficits, and psychosocial burden on families of afflicted children (e.g., Vallani et al., 2022).

There is evidence that OCD tends to manifest in some distinct ways in childhood versus adulthood. For example, whereas OCD occurs more frequently in males in youth compared to females (Geller et al., 2006), there is research showing that OCD is more common in females in adulthood (Fawcett, Power, & Fawcett, 2020). Additionally, earlier onset OCD has shown to be associated with greater overall symptom severity as well as the presence of comorbid mental health conditions when compared to OCD that has a later onset (Taylor, 2011). Given these unique features of OCD when it presents earlier in life as well as recent work showing that long-term treatment prognosis improves as a function of early recognition of symptoms (Liu et al., 2021), there is a need for effective assessments of OCD symptoms in youth.

There are a handful of assessment instruments that have generated good empirical support for measuring OCD symptoms in youth (Rapp et al., 2016). However, many of these assessments are hindered by methodological limitations that impact the clarity of understanding OCD symptoms in youth. To illustrate, most existing assessments of OCD in youth do not take avoidance behaviors into account, which is problematic in light of the highly prevalent nature of avoidance in individuals with OCD (Starcevic et al., 2011). Additionally, many of the available OCD symptom measures assess obsessions separately from compulsions, as though they are unrelated. However, research analyzing the structure of OCD symptoms shows that both obsessions and compulsions distill into dimensions based on functional relatedness (e.g., obsessional fears of being responsible for harm are related to checking compulsions; Deacon & Abramowitz, 2005). Finally, many existing measures of OCD tend to place greater emphasis on commonly-occurring symptoms of the condition (e.g., contamination-focused obsessions), which may lead to a confounding of symptom severity with symptom commonness. To illustrate, a child with contamination obsessions may appear, as reflected on the total score of a rating scale, more severe than a child with scrupulosity obsessions simply by virtue of a greater number of assessment items measuring contamination symptoms.

Based on the limitations in existing measures, there is a need to identify alternative assessments that can provide conceptual clarity about the nature and severity of OCD symptoms in youth. The Dimensional Obsessive Compulsive Scale (DOCS; Abramowitz et al., 2010) is a patient-rated scale that was developed with the specific aim of addressing limitations in extant measures of OCD symptoms; most significantly, it separately measures severity of four symptom subtypes. Numerous studies using the DOCS have provided support for its psychometric suitability in assessing OCD symptoms in adults and its subsequent translation and use in other languages and cultures around the world (e.g., Enander et al., 2012). However, despite the DOCS representing an improvement over alternative measures of OCD symptoms, to date there has been no examination of the DOCS in a youth sample.

The purpose of the present study was to examine the psychometric properties of the DOCS in a youth sample, including confirmation of the original factor structure, and convergent validity and sensitivity to treatment effects compared to another OCD scale designed for measuring symptoms in youth, the Obsessive Compulsive Inventory-Children’s Version-Revised (OCI-CV-R) (Abramovitch et al., 2022). Youth patients seeking treatment for OCD with consent provided by their parent(s) or legal guardian(s) were given the opportunity to participate in this study. In addition to typical clinical assessment and treatment procedures of an online teletherapy service, participants completed the DOCS and a previously validated measure of OCD symptoms in youth at the same time points in order to assess correlates with the DOCS. In this paper, we report the results of this research on the psychometric performance of the DOCS and its apparent clinical utility in a large youth sample.

## Methods

### Participants and Recruitment

Individuals being treated in an online teletherapy service specializing in treatment of OCD were recruited for voluntary participation in the study. As part of the intake process where parents or legal guardians review and sign a clinical consent form for their child to receive psychotherapy services, they were provided an additional voluntary research consent form describing participation in this study. This additional consent form explained that, in addition to standard clinical assessment procedures of the teletherapy service, study participation would include completion of one additional assessment measure at three separate time points during the course of therapy. Youth (n = 182) whose parent(s) or legal guardian(s) completed the informed consent form and who were subsequently diagnosed with OCD during the clinical intake process were eligible to participate in the study. The OCI-CV-R was sent to the patient for completion at the same time as the DOCs, at baseline, week 3, and week 9. This study was approved by a university Institutional Review Board.

## Measures

### Dimensional Obsessive Compulsive Scale (DOCS)

The DOCS (Abramowitz et al., 2010) is a 20-item self-report measure of OCD symptom severity across four empirically-derived symptom dimensions: (1) contamination, (2) responsibility for harm, injury, or bad luck, (3) unacceptable obsessional thoughts, and (4) symmetry, completeness, and exactness. The DOCS has previously demonstrated strong psychometric performance in multiple adult samples (Kuckertz et al., 2021; Ong et al., 2020; Safak et al., 2018). In previous research, it has shown strong convergent validity with the Obsessive Compulsive Inventory-Revised (*r* = .69) and the Yale-Brown Obsessive Compulsive Scale (*r* = .54) (Abramowitz et al., 2010).

### Obsessive Compulsive Inventory-Children’s Version-Revised (OCI-CV-R)

The OCI-CV-R (Abramovitch et al., 2022) is an 18-item self-report measure of OCD symptom severity in children and adolescents. Each item represents a feature common to OCD (e.g., “*I’m upset by bad thoughts*”) to which the respondent indicates the frequency of their experience by choosing one of three options: never, sometimes, or always. In previous research, the OCI-CV-R has demonstrated acceptable convergent and discriminant validity as well as internal consistency (Abramovitch et al., 2022). In the present study, the OCI-CV-R was used as a comparison to evaluate the performance of the DOCS in a youth sample via observing correlations between the two across three time points.

### Diagnostic Interview for Anxiety, Mood, and Obsessive Compulsive and Related Neuropsychiatric Disorders (DIAMOND)

The DIAMOND is an empirically validated semi-structured diagnostic interview that was used to assess and confirm a diagnosis of OCD in all study participants (Tolin et al., 2018). Additionally, the DIAMOND includes a two-item clinician administered severity rating scale that was used in this study to have a clinician-rated assessment of participants’ OCD symptom severity. Using this scale, the clinician makes separate ratings of their client’s psychological distress and impairment in functioning associated with OCD symptoms on a scale ranging from 1 (Normal) to 7 (Extreme). The higher of the two ratings is taken as the total severity score.

### Procedure

As part of the usual clinical treatment procedures, all participants were treated with exposure and response prevention (ERP) therapy, an empirically supported treatment intervention for OCD in youth. Based on research supporting an intensive initial phase of ERP as predictive of an optimal treatment response (Gershkovich et al., 2021), all participants were encouraged by their therapist to complete two 60-minute therapy sessions per week for at least the first three weeks of treatment. After this time, the results of assessment measures were used to gauge treatment response and make recommendations about continuing versus reducing the frequency of therapy sessions. For all, parents were required to be present for the first two sessions. During these sessions, the therapists provided instructions for the rating scales. Parental involvement in additional sessions was determined collaboratively between therapists and parents/caregivers.

As part of the standard clinical assessment procedures of the online teletherapy service, children and their parent(s)/legal guardian(s) completed the DOCS at the outset of treatment (“baseline”) and then repeatedly every three weeks (i.e., week 3, week 6, week 9, etc.) throughout their course of treatment. In the present study, in addition to the standard clinical assessment procedures, participants completed the OCI-CV-R at three time points: baseline, week 3, and week 9. We chose these three time points for OCI-CV-R completion to correspond with baseline OCD symptom severity, initial response to therapy (week 3), and treatment response after completing an amount of treatment (between 9 and 18 sessions) that is consistent with a standard course of therapy (week 9). The order of administration of the measures was randomized. Participants were given Amazon gift cards for each set of completed DOCS and OCI-CV-R scales. Participants who completed both scales at baseline (only) were given $5, those completing both measures at two study time points were given a $10 Amazon gift card, and participants who completed both measures at all three study time points were given a $30 Amazon gift card.

### Data Analysis

We used a linear mixed model with time point as a fixed factor and patient as a random factor to determine whether treatment resulted in a significant reduction in symptoms as measured by both the DOCS and the OCI-CV-R. Next, we performed a *Χ^2^* test to determine whether the outcomes on the two measures were independent based on the proportions who met criteria for a “response” on each scale: at least 35% reduction on DOCS and at least 25% on the OCI-CV-R at week 9 compared with week 0. Correlation matrices were then generated to further assess convergent validity of the DOCS and OCI-CV. Finally, a confirmatory factor analysis was conducted to identify whether the same four-factor structure previously found in adults on the DOCS (Abramowitz et al., 2010) is also present in youth.

## Results

### Participants

Out of 3086 youth patients who initiated services with NOCD, 842 did not meet inclusion criteria and n=1237 declined to participate. Of the 1007 who were eligible and provided informed consent, 825 did not complete both the DOCS and the OCI-CV-R on at least the week 0 and week 9 timepoints, leaving 182 whose data were analyzed in the “response” *Χ^2^* analysis. There were 133 participants who completed both the DOCS and the OCI-CV-R at all three timepoints (n=49 [26.9%] were missing a midpoint measurement). The mean age of the sample was 13.16 (*SD* = 2.91) years, with ages ranging from 5 to 17. With regard to gender, 50.6% (*n* =92) of participants identified as female, 35.7% (*n* = 65) of participants identified as male, and 13.7% (*n* = 25) of participants indicated nonbinary or another gender non-conforming identity, or chose not to respond to an inquiry about gender identity (See Table 1).

**Table 1.**

| **Characteristic** | **n** | **%** | **Mean** | **SD** |
| --- | --- | --- | --- | --- |
| Age | 182 |  | 13.16 | 2.91 |
| Gender |  |  |  |  |
| Female | 92 | 50.55% |  |  |
| Male | 65 | 35.71% |  |  |
| Other<sup>1</sup> | 25 | 13.74% |  |  |
| Ethnicity |  |  |  |  |
| White | 123 | 67.58% |  |  |
| Asian, Asian American,<br>or Pacific Islander | 16 | 8.79% |  |  |
| Hispanic or Latino | 6 | 3.30% |  |  |
| Multiple | 23 | 12.64% |  |  |
| Black or African American | 5 | 2.75% |  |  |
| Other | 4 | 2.20% |  |  |
| Undisclosed | 5 | 2.75% |  |  |
| DIAMOND severity (baseline) | 182 |  | 4.30 | 0.81 |
| DOCS (baseline) | 182 |  | 28.59 | 13.54 |
| OCI-CV-R (baseline) | 182 |  | 16.34 | 7.28 |

<sup>1</sup>Includes nonbinary or another gender non-conforming identity, or chose not to respond
DIAMOND = Diagnostic Interview for Anxiety, Mood, and Obsessive Compulsive and Related Neuropsychiatric Disorders
DOCS = Dimensional Obsessive-Compulsive Scale
OCI-CV-R = Obsessive Compulsive Inventory-Children's Version-Revised

### Demographics and psychometrics Symptom change

Study participants experienced significant symptom reduction over time as measured by both the OCI-CV-R (F(1,314.1)=107.7, p<.001) and the DOCS (F(1, 314.9)=119.3, p<.001) from the linear mixed model analyses. From the *Χ^2^* test, the relationship between these variables was significant: *X*^2^ (1, *N*=182)=26.87, p<.001, indicating that the measures are not independent of one another. Sixty met response criteria for both, 75 met neither response criteria, 25 met DOCS criteria only, and 22 met OCI-CV-R criteria only. The odds ratio was 8.20 (95% CI 4.2-15.9), indicating that the measures are positively related. This suggests that the DOCS and the OCI-CV-R classified OCD symptom response similarly. Cronbach’s alpha at baseline was acceptable for both measures (DOCS α= .88 OCI-CV-R α=.86).

### Correlations between DOCS and OCI-CV-R

To further assess convergent validity of the DOCS and the OCI-CV-R in this sample, correlations between the DOCS and the OCI-CV-R were calculated at baseline, 3 week follow-up, and 9 week follow-up. Strong positive correlations between the DOCS and the OCI-CV-R at baseline (r=.79, p<.001), 3 week follow-up (r=.80, p<.001), and 9 week follow-up (r=.81, p<.001), provide evidence for convergent validity.

Correlation matrices were generated to assess convergence of DOCS and OCI-CV-R subscales at all time points (Figure 2). Most notably, The DOCS contamination subscale was strongly positively correlated with the washing subscale of the OCI-CV-R at baseline (r=.84, p<.001), 3 weeks (r=.88, p<.001), and 9 weeks (r=.82, p<.001). Additionally, the DOCS unacceptable thoughts subscale was highly positively correlated with the obsessing subscale of the OCI-CV-R at baseline (r=.73, p<.001), 3 weeks (r=.76, p<.001), and 9 weeks (r=.73, p<.001).

**Figure 1.**
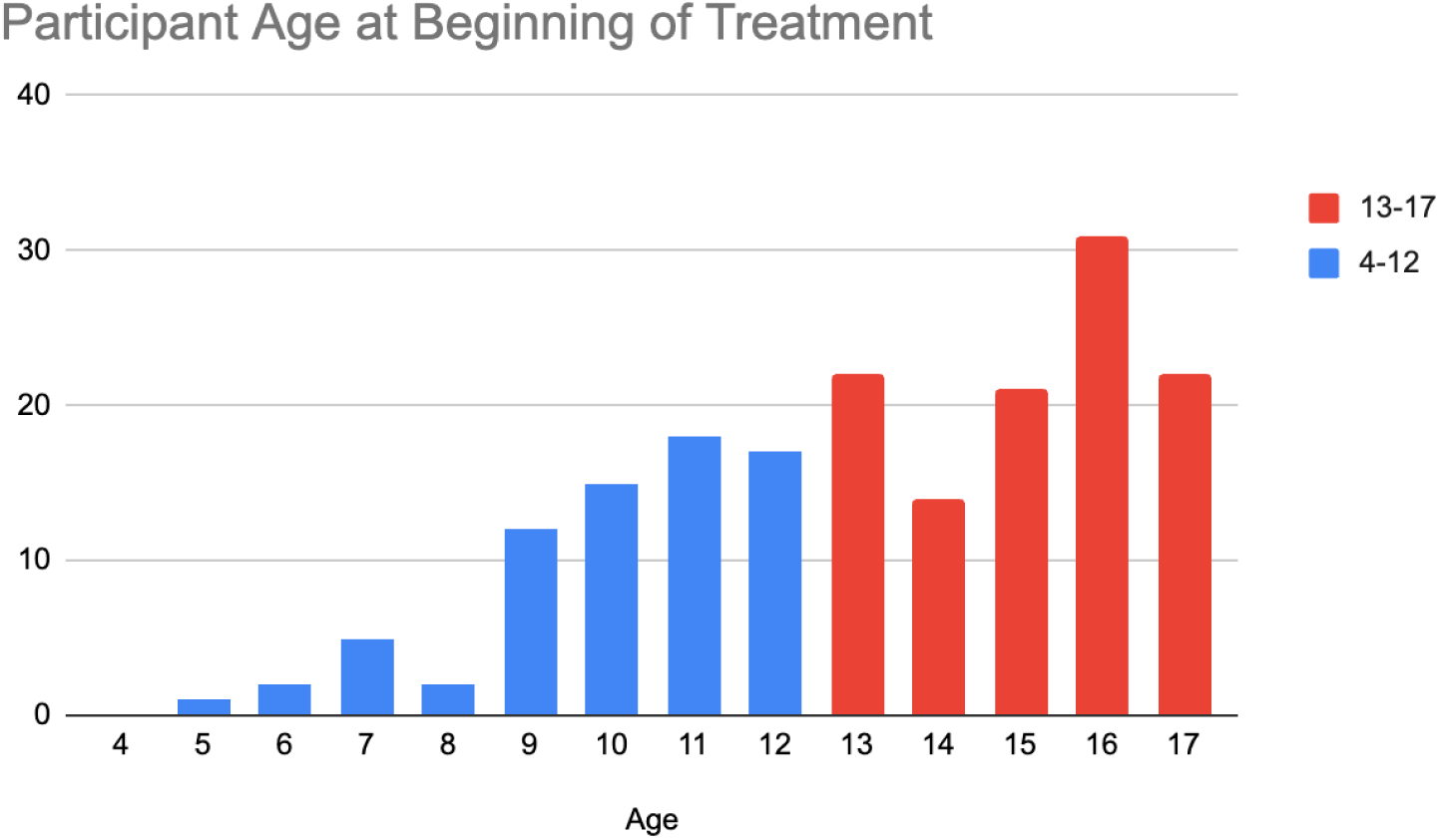
Age Distribution for the total sample

**Figure 2.**
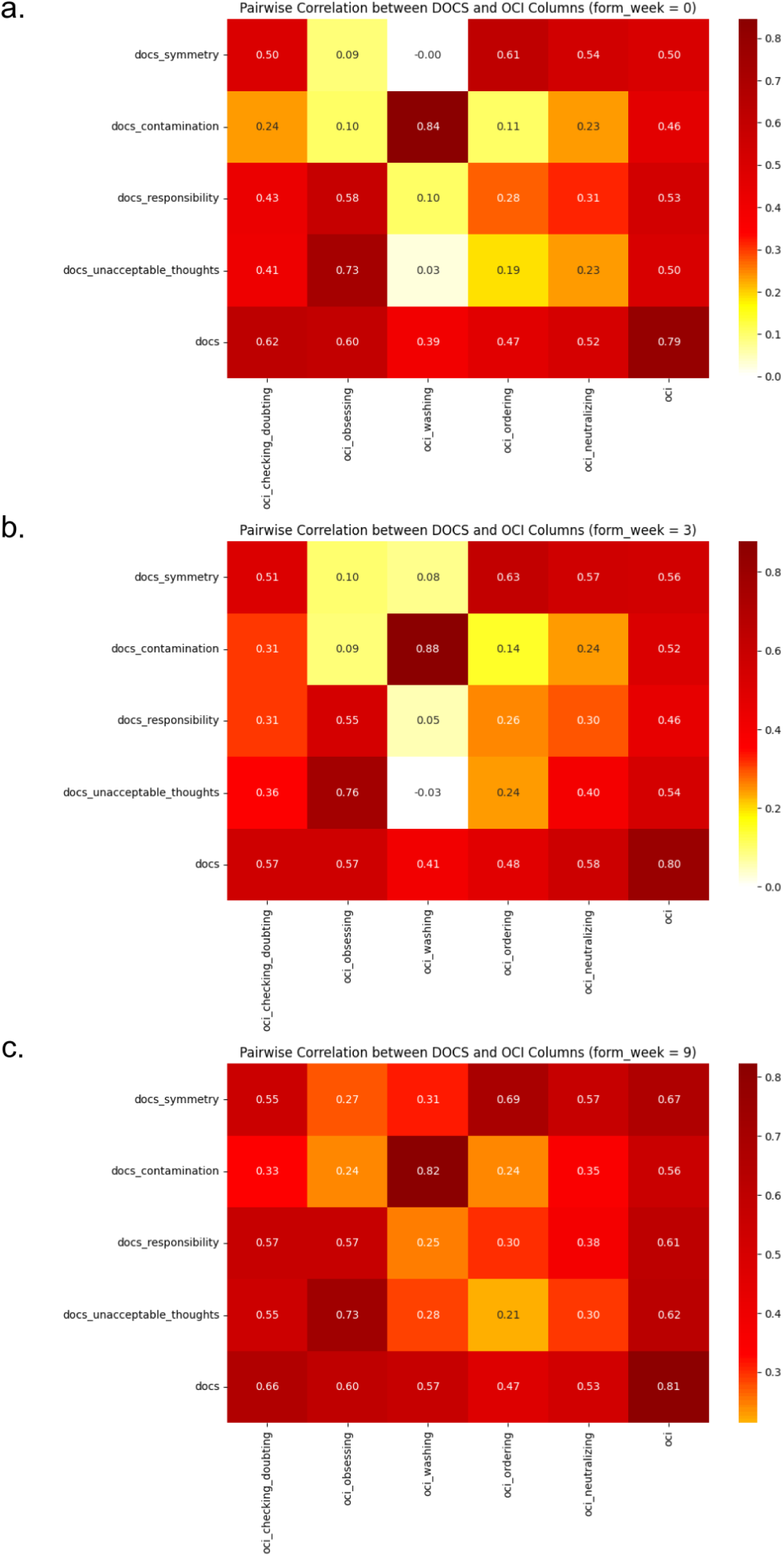
Pairwise Pearson correlations between DOCS and OCI-CV-R subscales. 2a shows the correlations at week 0 (N=182), 2b shows the correlations at week 3 (N=133), and 2c shows the correlations at week 9 (N=182). The bottom-right cell shows the correlation between total scores on the DOCS and OCI-CV-R. Darker shades of red denote stronger correlations between subscales, as indicated by the color bars on the right.

### Factor analyses

We conducted a confirmatory factor analysis (CFA) to identify whether the same factor structure is present for the DOCS in both adults and children. As in Ambramowitz et al. (2010), the goodness-of-fit indices supported the fit of the data to the four-factor model: RMSEA = .078, SRMR = .037, TLI = .937, and CFI = .945. Inspection of standardized residuals indicated no localized points of ill fit in the solution. Correlations among the four factors from the CFA (Table 2) showed a similar pattern as in adults (Abramowitz et al., 2010).

**Table 2.**
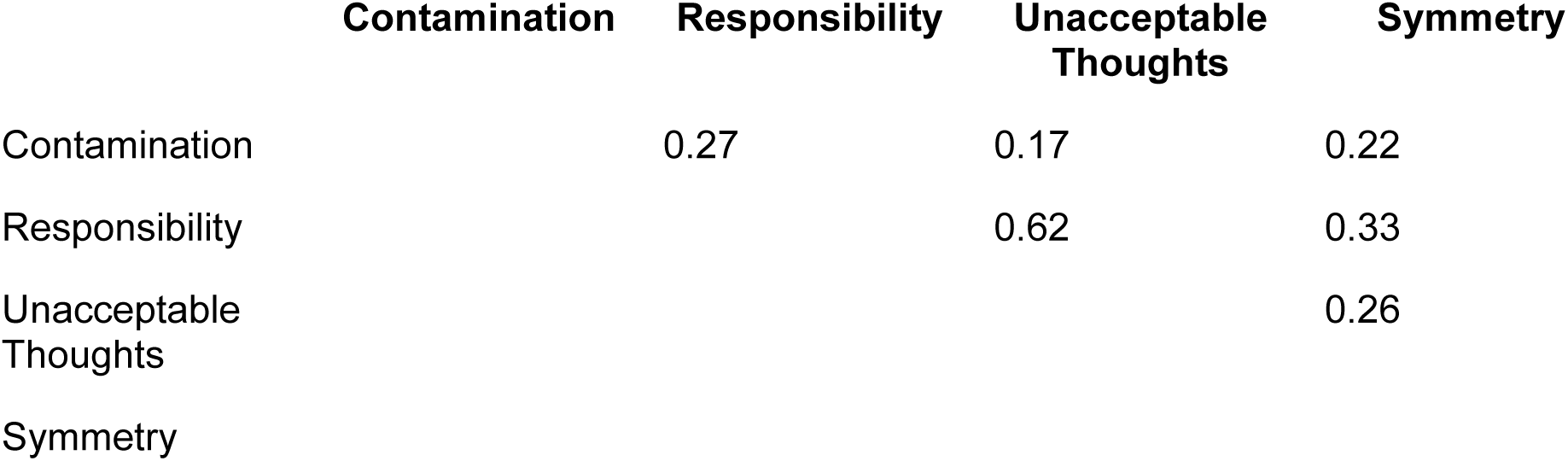
Correlations among the factors in the CFA of the DOCS.

|  | Contamination | Responsibility | Unacceptable Thoughts | Symmetry |
| --- | --- | --- | --- | --- |
| Contamination |  | 0.27 | 0.17 | 0.22 |
| Responsibility |  |  | 0.62 | 0.33 |
| Unacceptable Thoughts |  |  |  | 0.26 |
| Symmetry |  |  |  |  |

Next, we tested a higher-order CFA model to determine whether a single higher-order factor accounted for the interrelationships between the four lower-order factors. Each of the first-order factors loaded significantly on the higher-order factor, suggesting that the higher-order model provided a good account for the correlations among the first-order factors (Figure 3).

**Figure 3.**
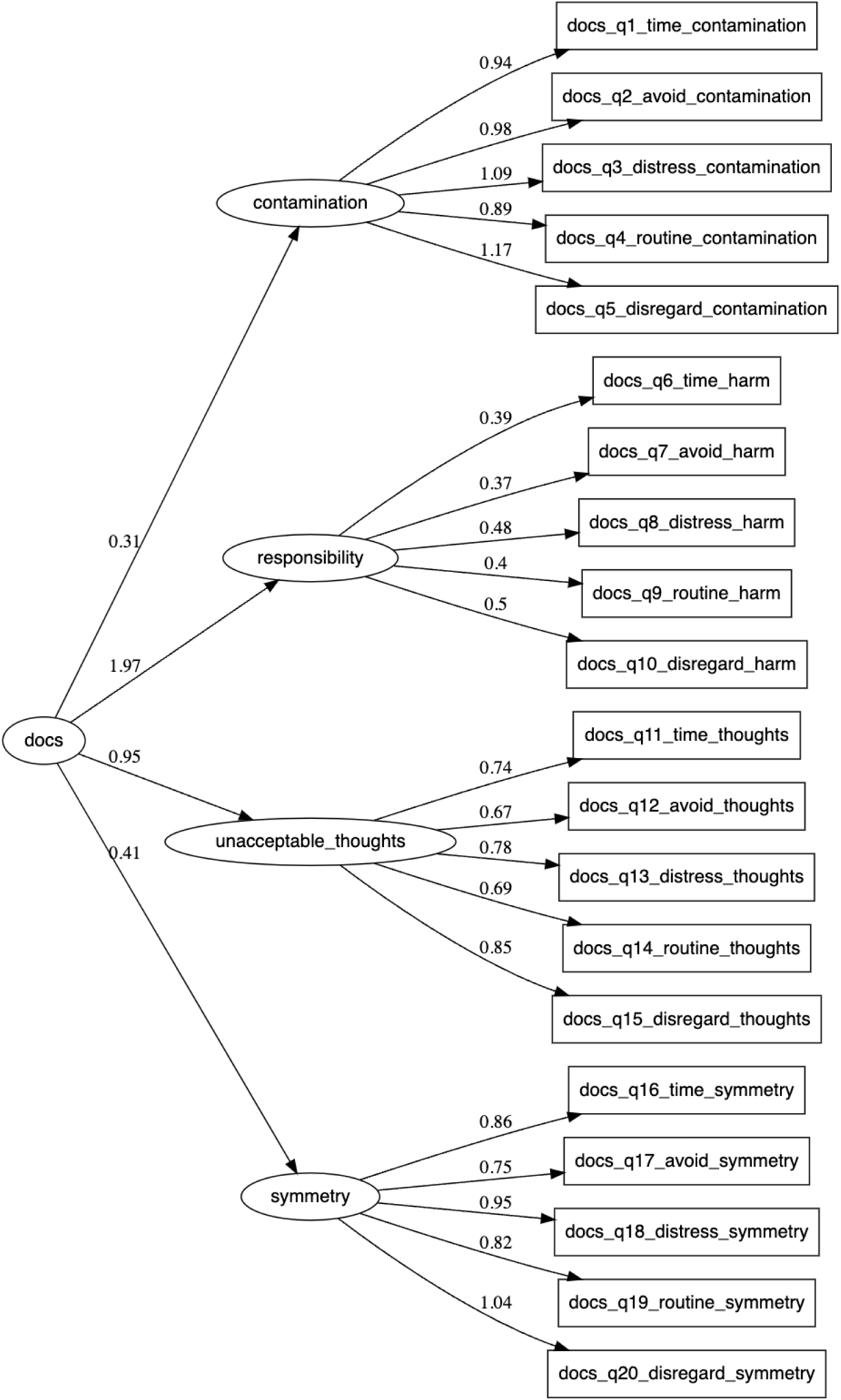
Path diagram with CFA results

The factor analyses confirmed the four-factor structure of the DOCS, with each factor representing a distinct dimension of obsessive-compulsive symptoms. The higher-order CFA indicated that these dimensions are part of a broader obsessive-compulsive construct. The factor loadings were strong, indicating that the items are good indicators of their respective dimensions. The model fit indices suggest that the DOCS is a robust measure for assessing obsessive-compulsive symptoms in youth, consistent with previous research in adult samples.

## Discussion

The goal of this study was to examine the psychometric properties of the DOCS in a youth sample and compare it to an instrument previously validated in this population, the OCI-CV-R. The relationship between the DOCS and the OCI-CV-R was examined across three time points during the course of ERP treatment for OCD among children and adolescents. Our findings demonstrate strong correlations between these two scales, supporting the relevance of using the DOCS in children and adolescents throughout treatment for OCD to identify and monitor symptom severity.

Participants in this study experienced reduced symptom severity during treatment, which was seen similarly on the DOCS and the OCI-CV-R. The significant symptom reductions seen through both of these scales suggests that they are examining similar trends in symptom change, as verified by the *Χ^2^* test. Moreover, significant positive correlations between the two scales remained similar at baseline, week 3, and week 9, even as symptoms were changing. In all, this suggests that the DOCS can satisfactorily measure cross-sectional symptom severity and change in OCD symptoms in children and adolescents with similar precision as the OCI-CV-R.

Through completing analysis of the subscales of these two measures, additional correlations were found to further support the association between these two measures. Most significantly, these associations were noticed between the OCI-CV-R obsessing subscale and the DOCS unacceptable thoughts subscale. Significant correlations were also found between the OCI-CV-R washing subscale and the DOCS contamination subscale. This highlights that both of these scales are able to identify similar markers of OCD subsets.

The confirmatory factor analysis supported the four factor model for the DOCS, which is consistent with findings found in adults in Abramowitz et al. (2010). The analysis indicated a good fit with the data and supports the idea that various dimensions of obsessive-compulsive symptoms are part of a broader construct. This further validates the DOCS as a comprehensive measure of OCD symptoms in children and adolescents with OCD, as well as adults (Enander et al., 2012).

Despite the strengths of this study, there are a few limitations. First, 27% of the participants did not complete both scales at the 3 week follow-up, which slightly reduces the confidence of the correlations observed at that timepoint. Additionally, because this was a naturalistic study in a real-world, treatment-seeking population and no standardized methods for completing the self-rated assessment scales were imposed, we cannot ascertain the extent to which parents or other caregivers may have been involved in the completion of the assessment measures. Indeed, the DOCS was initially authored for use with adults, and younger children may have had some difficulty understanding the wording of certain items on the DOCS and requested assistance from a parent/caregiver. Also, a surprisingly high number of prospective participants elected not to participate in the study, and it may be that there are characteristics among the study participants that set them apart from those who declined participation, which raises questions about the generalizability of this study’s findings to the population as a whole. Lastly, because the DOCS and the OCI-CV-R were presented together at each of the three administration time points (although randomized as to which was given first), it may be that completion of one scale influenced responses given on the other. Future research comparing the psychometric properties of the DOCS may consider administering it with further time separation from the other assessment measure.

## Conclusion

This study demonstrated the psychometric utility of the DOCS as a valid assessment of OCD symptoms in child and adolescent patients with OCD. DOCS total scores and subscale scores were consistently and significantly correlated with the total scores and subscale scores of the OCI-CV-R, a measure that has shown previous utility in assessing OCD symptoms in youth. Furthermore, the DOCS demonstrated appropriate sensitivity to change in a clinical treatment setting, which supports its consistent use throughout a course of treatment for OCD to measure responsiveness to treatment effects. Given the important contributions the DOCS has made to assessing OCD symptoms in a more dimensional manner, the results of this study support the use of the DOCS with children and adolescents with OCD in clinical and research settings.

## Data Availability

All data produced in the present study are available upon reasonable request to the authors

